# Mental health impacts of the COVID-19 pandemic on children and adolescents with chronic health conditions

**DOI:** 10.1101/2021.08.10.21261816

**Authors:** Louise M Crowe, Cathriona Clarke, Stephen Hearps, Remy Pugh, Nicky Kilpatrick, Emma Branson, Jonathan M Payne, Kristina M Haebich, Natalie McCloughan, Christopher Kintakas, Genevieve Charles, Misel Trajanovska, Ivy Hsieh, Penelope L Hartmann, Sebastian King, Nicholas Anderson, Vicki A Anderson

## Abstract

**Objective:** To investigate the changes in mental health and activities of children with chronic health conditions (CHC) during the pandemic. Additionally, to gather information from parents about their children’s healthcare experience, family stressors and mental health during the COVID-19 pandemic.

**Design:** A prospective longitudinal single site cohort study

**Setting:** Royal Children’s Hospital, Melbourne, Australia

**Participants:** 151 parents of children aged 1.5-17 years (M = 9.8 years, 58.3% male) with a CHC (colorectal disorder, cleft palate and neurofibromatosis type 1) completed the survey.

**Main outcome measures:** An adapted version of the COVID-19 Wellbeing and Mental Health Survey for Children and Adolescents (Parent/Caregiver version) (CRISIS) was utilised. The CRISIS tool provides data on child’s activities and mental health and parent mental health prior to, and during, the COVID-19 pandemic. Healthcare experiences families was also examined. Parents completed the Kessler 10.

**Results:** Compared to pre-COVID lockdown, parents reported their children were experiencing higher rates of loneliness, irritability, worry and anxiety. Parents reported that the restrictions in face-to-face contact with family and friends had been stressful for 80.0% of children. Children’s activities changed considerably during the COVID-19 pandemic with screen time increasing by 40%. Thirty percent of parents reported significant distress of their own. Parents felt telehealth were of poorer quality than face-to-face appointments.

**Conclusions:** Children with CHC experienced a significant increase in mental health symptoms during lockdown for the COVID-19 pandemic. Our findings highlight the increased need for clinical monitoring for children with CHC during periods of community stress and restrictions.

---

The coronavirus 2019 (COVID-19) pandemic has led to profound impacts on the lives of Australian children, particularly those in the state of Victoria where a state of disaster and stringent lockdown was in place for 18 weeks. Melbourne, Victoria’s capital city, has a population of approximately 5 million people (1). Its children have experienced major upheaval to their daily activities including remote learning, closure of playgrounds, playcentres, cultural and recreation centres (i.e., museums, zoos, cinema, etc), cancellation of organized sports and activities (i.e., music lessons, dancing), and restrictions on socializing with extended family and friends. For their families, and the community in general, stay-at-home orders were in place. At the highest level of restriction, which lasted over 3 months, families were only able to leave their home for one hour a day for exercise, and one family member could leave the house once a day to shop for essentials such as groceries and medication. Travel was restricted to a 5km radius from home and no visitors were allowed in the home, resulting in no face-to-face interactions with extended family, such as grandparents, babysitters or friends (2). Unemployment rates escalated, causing acute financial stress for many.

There is a shared assumption that this huge upheaval will have significant negative impacts on child mental health and wellbeing (3, 4). Behavioural and attentional difficulties in children have been reported at the times of peak restrictions (5). For children with chronic health conditions (CHC), who are already at elevated risk of mental health problems, the experience of the COVID-19 pandemic is likely to be magnified. These children represent a significant percentage of the patient population attending The Royal Children’s Hospital (RCH), the major tertiary paediatric hospital in Melbourne, servicing the state of Victoria. Compared to healthy children, children with CHC, and their families, have experienced additional stressors associated with the inability to access routine clinical care due to lockdown restrictions: loss of medical support and services and reduced access to home-based health services (i.e. telehealth for medical and allied health appointments). Therefore, we anticipated that the effects of the COVID-19 pandemic will be magnified for children with CHC, resulting in higher levels of psychological symptoms when compared with healthy children (6). For example, children with pre-existing respiratory problems had an increase in mental health symptoms during the COVID-19 pandemic (7), and this was also seen for children with special educational needs and neurodevelopmental disorders (5). Research from recent natural disasters, such as Hurricane Katrina, have shown that children with CHC demonstrate negative psychological consequences, including behavioral change and increased reporting of sadness and withdrawal (8).

Additional stressors associated with the COVID-19 pandemic, such as increase in unemployment, housing insecurity, and increased stress of parents who have been asked to work from home and support their children with home learning, are likely to further impact these already vulnerable children. Research has demonstrated that during the COVID-19 pandemic parents have had an increase in stress and feelings of caregiver burden (9). Having a child with a CHC is associated with higher levels of stress and physical symptoms related to stress (e.g., stomachaches) than parents of healthy children during the COVID-19 pandemic (10). Similarly, a recent report found that parents of children with additional needs experienced elevated stress levels in relation to almost all potential stressors surveyed (5). Given the limited literature available, this research on the mental health of children with CHC living through the pandemic is of significant importance.

The purpose of this study was to investigate the impact of the COVID-19 pandemic on children with CHC, as rated by their parent/caregiver. We investigated how children’s activities changed, whether their healthcare needs were being met, any change in mental health symptoms, sources of family stress, and the parent mental health. This study represents an important opportunity to understand adjustment to a global pandemic for children and families affected by CHC.

## Methods

This paper reported on wave 1 of a prospective longitudinal cohort project ‘COVID Resilience’ conducted at a single site. Wave 1 data were collected during the Vicotrian lockdown period and represent acute responses to the pandemic. Additional data will be collected at 3 and 6 months post baseline. Ethical approval was granted by the RCH Human Research Ethics Committee (HREC 64840).

Inclusion criteria for the CHC arm of the project were:

i. 0 to 18 years-old
ii. current patient of RCH
iii. diagnosis of chronic illness (colorectal disorders, neurofibromatosis type 1 [NF1], cleft palate) and have accessed an outpatient appointment in the last 12-24 months or on a current waitlist
iv. parent/caregiver has sufficient English to complete questionnaires.

Participants were excluded they had a progressively deteriorating conditions (e.g., palliative or waiting for transplant) or if they had been an inpatient at RCH during the lockdown period.

### Measures

The surveys were delivered online via REDCap (11). The *COVID-19 Wellbeing and Mental Health Survey for Children and Adolescents (Parent/Caregiver version*) is a modified version of the CoRonavIruS Health Impact Survey (CRISIS) tool, which was developed to measure the psychosocial impact of the COVID-19 pandemic on individuals across mental, behavioural and physical health domains (12). The questionnaire captures pre-existing risk and protective factors, as well as health outcomes and behaviours following the pandemic (13). The current study adapted the CRISIS tool to seek specific information to our site, including health utilisation data. One parent/caregiver was asked to complete the CRISIS tool, comparing activities and symptoms for the three months prior to the pandemic with those for the past month (i.e. during lockdown). Symptoms were rated using a 5-point Likert scale, from not at all (1) to extremely (5).

The *Kessler 10 (K10)* (14) was completed by the child’s parent/caregiver. It is a 10-item adult measure used to assess mental health and psychological distress, with a clinical cut-off > 20. Scores of 20-24 indicates mild distress, 25-29 indicates moderate distress, and >30 indicates severe distress.

### Procedure

Clinicians and research assistants screened patients for eligibility using the hospital’s electronic medical record (EMR). Parents of children who met the inclusion criteria were sent an invitation letter, with details about the study, from their respective RCH clinical department. Families who did not opt out were contacted by phone to further explain the study, obtain informed consent and collect contact details. Once enrolled, one parent completed the baseline questionnaires online using a REDCap database. Upon completion of the baseline questionnaires, participants will be sent follow up surveys at 3-months (T1) and 6-months (T2) from their enrolment date to capture longitudinal information.

### Statistical analysis

Data are presented with descriptive statistics including percentages for categorical variables, and mean (and standard deviation [SD]) for continuous variables. Paired samples *t*-tests were used to compare child mental health symptoms pre-COVID and during COVID, with Cohen’s d reported as a measure of effect size (15). Wilcoxon signed-rank test was used to compare screen time hours (ordinal) over the two time points. All analyses were carried out using Stata 16.1, and employed a significance level of p < 0.05.

## Results

Data were collected between September and December 2020 and 297 families were contacted, 177 consented of which 151 families completing the survey. Due to ethics restrictions, we are unable to report why families decided not to complete the questionnaires. Table 1 lists the demographic information of the group.

**Table 1.** Group demographics.

|  | Total<br>n=151 |  | Cleft palate<br>n=65 |  | Colorectal<br>n=38 |  | NF1<br>n=48 |  | p |
| --- | --- | --- | --- | --- | --- | --- | --- | --- | --- |
| *Child characteristics* |  |  |  |  |  |  |  |  |  |
| Age [years], M (SD) | 9.8 | (4.1) | 9.4 | (3.6) | 8.7 | (3.9) | 11.3 | (4.5) | 0.005 |
| Sex [male], n (%) | 88 | (58.3) | 35 | (53.8) | 22 | (57.9) | 31 | (64.6) | 0.525 |
| Education setting, n (%) |  |  |  |  |  |  |  |  |  |
| Creche/daycare | 7 | (4.6) | 3 | (4.6) | 2 | (5.3) | 2 | (4.2) | 0.688 |
| Pre-school/kinder/ELC | 22 | (14.6) | 12 | (18.5) | 6 | (15.8) | 4 | (8.3) |  |
| Primary/Secondary school | 117 | (77.5) | 49 | (75.4) | 28 | (73.7) | 40 | (83.3) |  |
| Not attending any | 5 | (3.3) | 1 | (1.5) | 2 | (5.3) | 2 | (4.2) |  |
| *Parent characteristics* |  |  |  |  |  |  |  |  |  |
| Respondent, n (%) |  |  |  |  |  |  |  |  |  |
| Birth mother | 137 | (90.7) | 57 | (87.7) | 34 | (89.5) | 46 | (95.8) | 0.199 |
| Birth father | 9 | (6.0) | 5 | (7.7) | 3 | (7.9) | 1 | (2.1) |  |
| Adoptive mother | 3 | (2.0) | 3 | (4.6) | 0 | (0.0) | 0 | (0.0) |  |
| Foster father | 1 | (0.7) | 0 | (0.0) | 0 | (0.0) | 1 | (2.1) |  |
| Grandparent | 1 | (0.7) | 0 | (0.0) | 1 | (2.6) | 0 | (0.0) |  |
| Born in Australia, n (%) | 126 | (83.4) | 51 | (78.5) | 33 | (86.8) | 42 | (87.5) | 0.412 |
| Indigenous Australian, n (%) | 4 | (2.6) | 1 | (1.5) | 1 | (2.6) | 2 | (4.2) | 0.818 |
| English only, n (%) | 135 | (89.4) | 53 | (81.5) | 38 | (100.0) | 44 | (91.7) | 0.008 |
| Partnered, n (%) | 114 | (75.5) | 51 | (78.5) | 28 | (73.7) | 35 | (72.9) | 0.763 |
| Highest education, n (%) |  |  |  |  |  |  |  |  |  |
| Less than High School | 11 | (7.3) | 4 | (6.2) | 4 | (10.5) | 3 | (6.3) | 0.442 |
| High School | 15 | (9.9) | 4 | (6.2) | 4 | (10.5) | 7 | (14.6) |  |
| Certificate/Diploma | 52 | (34.4) | 22 | (33.8) | 11 | (28.9) | 19 | (39.6) |  |
| Bachelor's Degree | 38 | (25.2) | 15 | (23.1) | 10 | (26.3) | 13 | (27.1) |  |
| Postgraduate | 35 | (23.2) | 20 | (30.8) | 9 | (23.7) | 6 | (12.5) |  |

The mean age of 9.8 years translates to middle primary or elementary school age. Of the 117 children attending school, 72.6% (n = 85) were participating in remote learning. Parents were asked to indicate whether the amount of time their child participated in their usual (pre-COVID-19) activities had changed. Results indicated that children had spent more time on: computer games/screens/TV (60.3%); arts and crafts (52.3%); sports/active games (up by 44.4%); talking/discussion (76.2%) and creative play (39.1%).

Children spent significantly more time on screens during the pandemic than previously. Refer to Figure 1. Pre-COVID-19, only 9.9% of children were spending more than 4 hours a day on screens, however, during lockdown this percentage increased to 43.0% of children. A Wilcoxon signed-rank test showed the change in screen hours was statistically significant (*Z*=-9.00, *p* <.0001). Of note, this figure incorporates screen time for both for education and entertainment.

**Figure 1.**
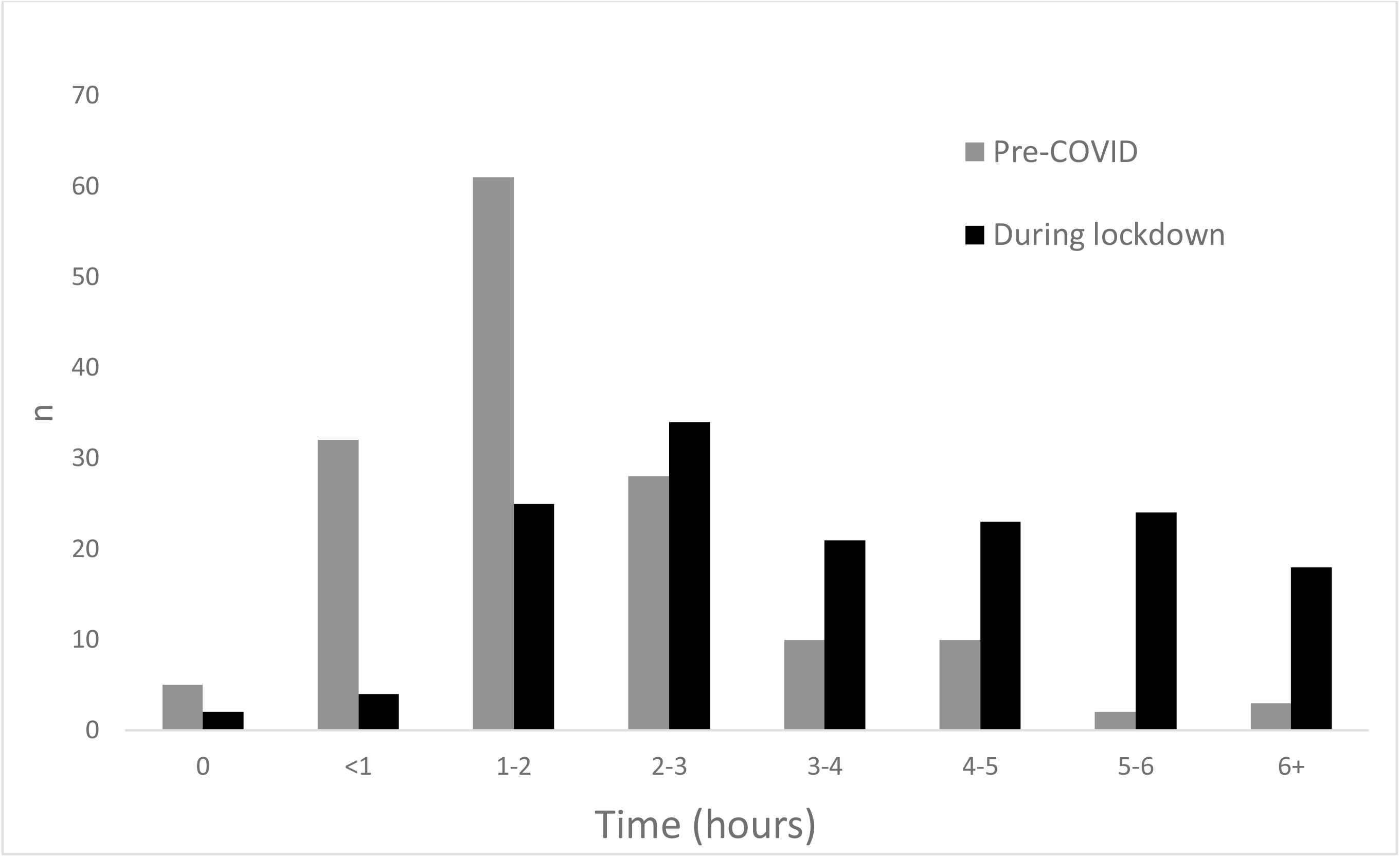
Time spent on screens by children pre-COVID and during lockdown. Figure 1 created by the author and used with permission.

### Children’s Healthcare Needs

Parents were asked about changes to their child’s healthcare needs, 55.6% (n = 84) indicated that their healthcare was unchanged, 31.1% (n= 47) indicated it was ‘a little worse’, and 6.6% (n = 10) indicated it was ‘much worse’. Just 6.6% (n= 10) felt their quality of healthcare had improved. Of those who had participated in telehealth, (n = 130), the majority at 54.6% indicated that their experience of telehealth was of worse quality than face-to-face care, 28.5% felt it was no different and 10.6% of families preferred telehealth over face-to-face appointments.

### Children’s Mental Health

Parents rated their child’s mental health symptoms for the 3 months pre-and during the pandemic lockdown, see Table 2. For each mental health symptom, parents reported a significant increase in symptoms in the COVID-19 pandemic compared to pre levels. Symptoms that increased the most included loneliness (Cohen’s *d* = 0.76), irritability (*d* = 0.62), worry (*d* = 0.62), and anxiety (*d* = 0.60). Parents reported that the changes in contacts with family and friends had been stressful for 80.0% of children.

**Table 2.** Severity of mental health symptoms pre-COVID-19 and during lockdown (n=151)

| Symptoms | Severity | Pre-COVID, n (%) | Past Month, n (%) |
| --- | --- | --- | --- |
| *Anxious\** | Moderately | 21 (13.9) | 38 (25.2) |
|  | Very | 2 (1.3) | 7 (4.6) |
| *Fatigue\** | Moderate-very | 32 (21.2) | 51 (33.8) |
|  | Extremely | 0 | 2 (1.3) |
| *Depressed/Unhappy\** | Moderately | 6 (4.0) | 24 (15.9) |
|  | Very | 0 | 1 (0.7) |
| *Irritable/ Easily angered* | Moderate-very | 45 (29.8) | 66 (43.7) |
|  | Extremely | 3 (2.0) | 15 (9.9) |
| *Lonely\** | Moderate-very | 23 (15.2) | 58 (38.4) |
|  | Extremely | 0 | 4 (2.7) |
| *Distracted* | Moderately | 38 (25.2) | 34 (22.5) |
|  | Very | 7 (4.6) | 15 (9.9) |
| *Worried\** | Moderate-Very | 28 (18.5) | 52 (34.4) |
|  | Extremely | 1 (0.7) | 4 (2.7) |

\*significant difference in symptom level pre-COVID-19 and during lockdown

### Family Stress

For those parents whose child was participating in remote learning (n = 85), when asked how difficult it was to balance remote learning with paid work demands, 64.7% (n = 55) of parents indicated it had been quite to very difficult, and 51.8% (n = 44) indicated it had been difficult to balance with other household responsibilities. When asked to identify any positive changes 37.2% (n = 35) reported that there had been some positive changes in their family and community, including ‘more quality time together as a family’ (85.7%, n = 30), followed by ‘less running around’ (71.4%, n = 25) and ‘people looking out for each other’ (48.6%, n = 17).

### Parent stress levels

Parents rated their stress level during the pandemic lockdown from 1 (not stressful) to 10 (very stressful). The mean response was 5.41 (SD= 2.45), with 14% (n=16) reporting that their relationship with their partner had deteriorated. The mean K10 score for the sample was 18.97 (SD = 7.10), which falls just below the clinical cut off for psychological distress of > 20. However, 25 parents (19.53%) had a score between 20 and 24 indicating mild distress, 12 (8.0%) parents had moderate distress, and 12 (8.0%) parents reported severe distress.

### Discussion

The purpose of this study was to investigate the impact of the COVID-19 pandemic on children affected by CHC and their families. Data related to children’s daily activities, healthcare experience, stress of families, and mental health of parents were collected via an online survey tool. Our preliminary findings confirm that strict and lengthy lockdown during the COVID-19 pandemic led to significant changes in the ways that children with CHC and their families lived their lives which, in turn, were associated with increases in child mental health symptoms and parent distress.

The majority of children were restricted to their homes during the lockdown and were required to participate in school remotely, with no face-to-face contact with peers. Many normal leisure pursuits and activities were also ceased. Given these restrictions, it is not surprising that parents reported significant increases in their children’s time playing computer games and watching screens and TV during the pandemic lockdown. Prior to the stay-at-home orders, most children were on devices for only 1-2 hours a day; however, during the lockdown, this increased to 2-3 hours day. Heavy use of devices (> 4 hours/day) increased more than fourfold from 9.9% to 43.0% of children during the pandemic. The addictive nature of devices and negative consequences is somewhat of a concern (16-17). Other activities including arts and crafts, home based sports and active games and creative play also all increased.

Children with CHC are typically linked to a hospital outpatient clinic where they attend regular face-to-face appointments to access their healthcare. During the extended lockdown experienced by Victorian children, appointments were moved to telehealth. While telehealth may be more accessible and its use may continue after the COVID-19 pandemic, the majority of families in this study reported telehealth to be of lower quality than face-to-face appointments. Parents indicated that they did not feel their healthcare needs were met as well overall during the COVID-19 pandemic likely due to reduced/changed access to therapies (e.g., physiotherapy, speech therapy), carers, and support workers (e.g., respite care, educational assistants). We are unaware of other research reporting the healthcare experience of children with CHC during the pandemic.

The mental health impact of the COVID-19 pandemic has been frequently reported in the media and is now emerging in the scientific literature, and was a particular focus of our study. For our participants, the greatest change was associated with loneliness, which is consistent with the limitations in socialisation with peers during lockdown. Of note, in keeping with prior research regarding social isolation of children with CHC, moderate levels of loneliness were identified in a small subset of children pre-pandemic, but this increased significantly in frequency and severity during the pandemic, likely due to a combination of missing interactions with extended family and friends.

There has been reported increases in reported loneliness during the COVID-19 pandemic in adolescents (18). In our sample, parents highlighted that social isolation was stressful for the majority of children during the COVID-19 restrictions. Of note, loneliness, irritability, worry and anxiousness increased significantly with COVID-19 restrictions, similar to findings after natural disasters (8). This is consistent with research showing a deterioration in mental health of children with respiratory disorders during the COVID-19 pandemic (7).

Despite changes in children’s mental health and wellbeing, most parents rated their own stress level at an average level, with 16% reporting moderate to severe distress. Parents also described difficulties meeting the demands of work due to additional childcare responsibilities, with similar rates to UK based research (5). Most parents identified positive aspects of stay-at-home orders, including increased quality time with family, less running around, and people looking out for each other. The uniqueness of the COVID-19 pandemic, with its ‘whole of society’ shared experience, may provide a buffer to the stress experienced, with a feeling of solidarity and community that ‘we are all in this together’.

Limitations to these findings include that we have captured pre-COVID-19 status in some areas, with the associated risk of parent retrospective reports reflecting the “good old days”. We did not include self-report, and so ratings of child function may be confounded by parent coping and stress.

Our study has identified that the COVID-19 pandemic has had multiple negative impacts on children with CHC and their families, spanning child activities, health services, and child mental health. Our findings support the need for clinicians to monitor the mental health status of children with CHC and consider the quality of telehealth delivery of clinical services.

## What is already known on this topic

- Behavioural and attentional difficulties in typically developing children were reported at the times of peak restrictions
- Little is known about the impact of the pandemic on children with chronic health conditions, however, children with pre-existing difficulties such as special educational needs and neurodevelopmental disorders had high levels of parent-reported mental health symptoms in the pandemic

## What this study adds

- This study represents an important opportunity to understand adjustment to a global pandemic for children and families affected by chronic health conditions.
- Findings include that children with chronic health conditions had significant increases in parent-rated mental health symptoms
- Parents felt that telehealth or online medical appointments were of poorer quality than face to face appointments

## Data Availability

Reasonable requests for data sharing will be considered.

